# Protocol for an individual-level, two arm, superiority RCT within an adaptive platform trial: Enhanced patient counselling and SMS reminder messages to improve access to community-based eye care services in Meru, Kenya

**DOI:** 10.1101/2024.02.28.24303254

**Authors:** Luke Allen, Min Kim, Michael Gichangi, David Macleod, James Carpenter, Malebogo Tlhajoane, Sarah Karanja, Nigel Bolster, Matthew Burton, Andrew Bastawrous

## Abstract

**Background:** The Vision Impact Project (VIP) is a major community-based eye screening programme running in Kenya with the aim of promoting eye health for all. Previous studies embedded within the programme in Meru County have found that a third of people who are screened require care for an eye problem, however only half of these people manage to access outreach treatment clinics. Access varies between sociodemographic groups, and only 30% of young adults (18-44 years old) were able to access care. In previous mixed-methods work our team conducted interviews and surveys with non-attenders from this ‘left-behind’ group to explore what could be done to improve access.

**Methods:** Younger adults told us that better counselling at the point of referral would be likely to improve attendance rates. Based on their feedback, we have developed a script that will be read to participants in the intervention arm at the point of referral, and then sent as a reminder SMS the following day. We will assess whether attendance rates are higher among those randomised to receive this enhanced counselling compared to those who receive standard care. The primary outcome will be the proportion of people from the left-behind group who attend triage clinic. Our secondary analysis will examine overall mean attendance across all groups. We will calculate Bayesian posterior probabilities of attendance in each arm every seven days and continually recruit participants until one of two stopping rules have been met: there is a >95% probability that one arm is best or there is a >95% probability that the difference between the arms is <1%.

**Discussion:** This Bayesian RCT will be embedded into the clinical workflow software that is used to manage referrals and clinic attendance. It will test whether a simple, low-cost, service user-derived intervention is able to improve access to services among a population group that is currently being left behind.

**Trial Registration:** ISRCTN <u>11329596</u>, Registered on 02 February 2024

**Administrative Information:** 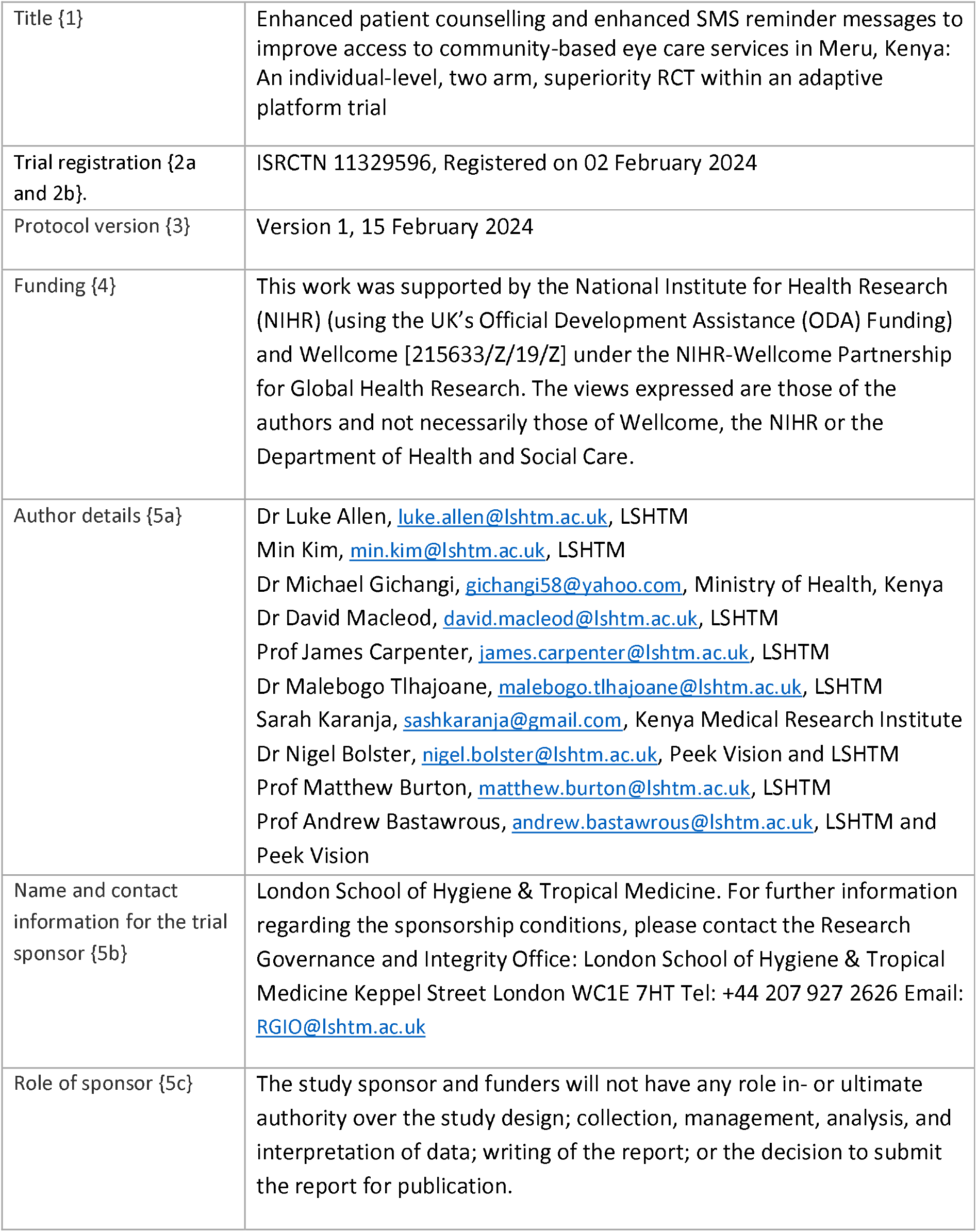

## Introduction

### Background and rationale {6a}

Approximately 1.1 billion people (over 10% of the global population) live with vision impairment that could be easily corrected.^10^ Two very cost-effective interventions - spectacles and cataract surgery – could eliminate over 90% of all vision impairment worldwide. Although provision of these services has risen in recent decades, effective coverage rates exhibit marked socioeconomic gradients at the international and intra-national levels, for example, the global effective refractive coverage is reported at 36%, with high-income countries reporting 90% and low-income only 6%.^10^

In major eye screening programmes, once people have been identified with an eye need and referred on, only around 30-50% of these people access care, and research from Nigeria and Sri Lanka suggests that unmarried (primarily widowed) women and people living in rural areas are the least likely to access care.^11^

This protocol outlines an intervention to be implemented in Kenya’s Vision Impact Project in Meru County. Previous studies conducted by our team found that only 50% of people found to have an eye care need during screening were able to access local treatment outreach clinics, once referred. An equity analysis found that age was associated with access: only 30% of younger adults (aged 18-44 years) accessed care once referred.

In interviews with younger adults who were not able to access care we identified a number of barriers and potential solutions to improve access to care for this group. We then conducted a survey with 401 additional young adults who were not able to access care and asked them to rank the potential solutions/service modifications by likely impact. One of the top-rated ideas was to provide additional information about treatment outreach clinics at the point of referral and in follow-on SMS reminder messages. Specifically, younger adults who were not able to access care told us that enhanced counselling should include information on;

- The outreach treatment clinic opening times,
- the services that are available at these clinics (vs those that require onward referral to hospital-based services),
- any costs involved at the outreach clinic,
- and the importance of attending

This trial is intended to test whether provision of this additional information is associated in a higher probability of accessing care. The trial is being conducted under an overarching adaptive platform trial protocol that is being used to test multiple low-risk service modifications to improve access to care, with a focus on ‘left-behind’ groups.

### Objectives {7}

To test whether provision of additional information around clinic opening times, services, costs, and the important of attending via in-person counselling at the point of referral and via reminder SMS messages increases the probability of accessing treatment outreach clinics compared to standard care.

### Trial Design {8}

This is a Bayesian, pragmatic, superiority, two-arm, individual-level, randomised controlled trial, embedded within the Vision Impact Project screening programme in Meru, Kenya. We will use routinely collected referral and attendance outcome data derived from the patient management and flow software.

## Methods

### Study setting {9}

This trial will be embedded within the Vision Impact Project (VIP) that is operating in Meru, Kenya. The programme has screened over one million people in the past year using a simple smartphone-based visual acuity screening app. Hundreds of thousands of people have been identified with an eye need and referred for free further assessment at local treatment outreach clinics. However, only half of those referred have been able to access this free care.

Our trial will be integrated into the screening and patient management software developed by Peek Vision. Peek Vision is a leading provider of eye screening software worldwide. The ‘Peek Capture’ app is used to screen participants for vision impairment, to capture observations by screeners and health practitioners, and to gather demographic data, as well as linking participants to a referral system that tracks each of their progression through the local eye health system. The same app is used to collect data on visual acuity, socioeconomic status, referral status, and attendance status (our primary outcome). Our trial will use these routinely collected data to test whether a series of interventions are able to reduce the proportion of people from marginalised groups with an eye care need who do not attend triage clinic once referred.

### Eligibility criteria {10}

As a pragmatic trial, the eligibility criteria are determined by the local VIP programme. We will include all adults (>18 years) who participate. We will exclude those who do not meet local clinical service eligibility criteria.

### Consent {26a, 26b}

Informed consent will be sought by screeners during screening - at the point that participants are identified as having an eye care need and referred on for further care. At the time of consenting, participants will receive detailed information about the research project including the objectives and measures taken to respect the confidentiality of the data collected. Consent will be recorded digitally using an electronic tick box (as appropriate for low-risk trials). The consenting process and the provision of participant information will be delivered through EpiCollect, a mobile phone data gathering tool with an associated web application, providing two-way communication between multiple data gatherers and a project database. This platform will be used solely for the digital consenting process and will be used alongside the Peek Capture App that is used during screening. Participants will be given the contact details of the research managers and will be free to leave the trial at any time. There will be no remuneration for participants.

### Interventions

#### Interventions and administration {6b, 11a}

The intervention is a script and reminder SMS message that have been developed in line with suggestions from intended service beneficiaries from the left-behind group. During interviews with 67 non-attenders from the left-behind group, 25 different potential service modifications were suggested. We then asked 401 additional non-attenders from the left-behind group to ascribe a simple score to each suggestion, ranging from ‘likely to make a large difference’ to ‘likely to make a small/no difference’ on a three-point Likert scale. The top-ranked suggestions were discussed at a workshop with representation from the VIP programme, the programme funder, the programme implementing partner, the county health management team, and the community advisory board. This group unanimously agreed that it would be feasible to implement and test a counselling intervention that bundled together four suggestions: that those referred be informed of the treatment outreach clinic opening times, the services that are available at these clinics (vs those that require onward referral to hospital-based services), any costs involved at the outreach clinic, and the importance of attending. A draft script that included these elements was reviewed and revised by all of the above stakeholders and a lay representative/intended service beneficiary from the left behind group. The text was translated into Swahili and back-translated into English to check that meaning had not been lost.

> **Control arm: usual care referral counselling**
>
> *“I have examined your eyes, and you have a problem, I have referred you in the system and you will receive an SMS with where and when you are supposed to attend treatment. You will come for treatment on [Date] at [location], the examination will be free and you will be informed of anything else on the material day*.
>
> **Intervention arm: enhanced referral counselling script**
>
> *“I have found a problem with your eyes. I am referring you to the outreach treatment clinic that will be held at [location] on [date] between [time] and [time]. At the clinic, eye care professionals will perform a specialist assessment and provide medicines and spectacles as required, all free of charge. Note that a small proportion of people who attend the clinic will be found to have complex eye problems that require onward referral for hospital assessment and specialist glasses. This may incur a cost. However, the vast majority of people have their needs fully met for free at the outreach triage clinic and do not require any further referral*.
>
> *With treatment, you will be able to see more clearly. This will help with your work, reading, viewing screens, and many other things. It is important that you attend the clinic or your eye problem may get worse. The clinic will only be running from [day] to [day], so if you don’t manage to attend, you may not be able to get free care again in the future*.*”*

The relevant script will be read out to the participant by the screener at the point of referral. The wording of the usual care counselling script is based on the screening programme training materials and observations of what screeners currently tell participants. No elements have been removed i.e. this script accurately reflects usual care. Screeners do not usually read this information out to participants; however, we are introducing standardised wording to reduce the risk of contamination i.e. screeners delivering the same enhanced counselling elements to participants in both the intervention and control arms.

All people who are referred are sent automated SMS reminder messages on the day of referral, the day before the appointment, and on the appointment day. These messages are generated and sent by the Peek Vision app. The content of the intervention SMS was developed by the research team in collaboration with a lay representative from the left-behind group. The messages are sent in either English or Kiswahili, depending on the participant’s chosen language.

> **Control SMS Script**
>
> *Dear <<name>>, you were examined and found to have an eye problem. Kindly report on <<location>> on <<date>> for assessment. For more information contact Meru Referral Hospital*.
>
> **Intervention SMS script**
>
> *We found that you had an eye problem. Please attend the outreach clinic at <<location>> on <<date>> between 9am-5pm. The specialist assessment is free If you are found to have a complex eye problem, you may be referred to a hospital for further care or specialist glasses, and this may include a fee*
>
> *However, the vast majority of people who attend the outreach get their eye problem fixed without the need for any further referral*
>
> *It’s important that you attend, as your eye problem may get worse, and you might not have a future opportunity to access free care. See you on <<day>>*

Figure 1 shows the point at which the interventions are delivered.

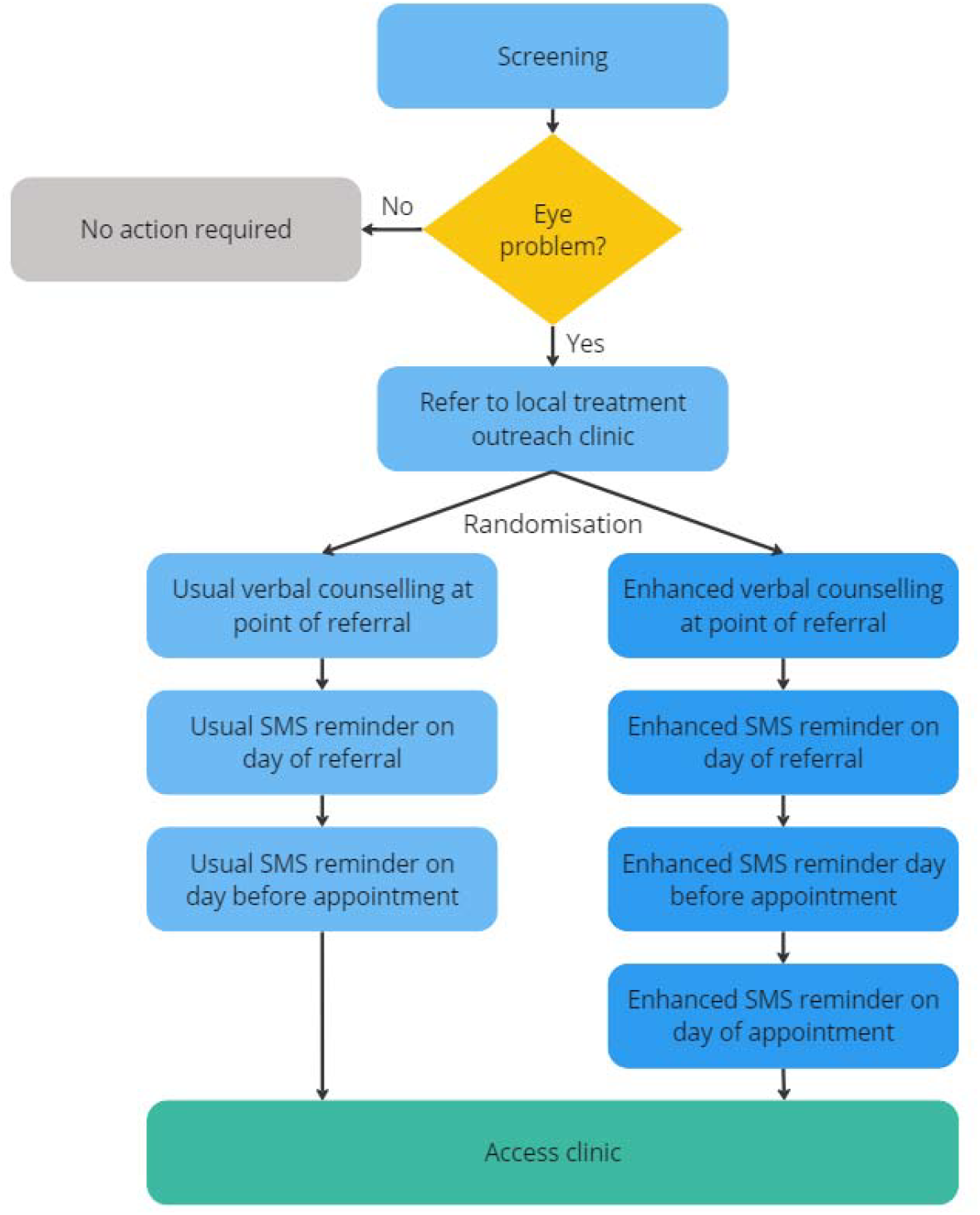

The theory of change is based around a classical information deficit model of behaviour change. The intervention is needed because many people are not accessing services, and these people tell us that an important barrier is the lack of clear information about opening times, services offered, costs, and the importance of attending. We will provide this information verbally at the point of referral and send a summary via SMS to those in the intervention arm. We will test whether those who receive this enhanced counselling information are more likely to attend than those who do not.

Whilst the information deficit model has received justifiable criticism for oversimplifying behaviour change - often in the context of paternalistic uninvited information provision^16^ - this intervention was suggested by people who told us that they genuinely could not access services for want of basic information. The wider literature suggests that SMS reminders can play a small but important role in improving access to care,^12–14^ however there is much less research on the provision of verbal information at the point of referral.^15^

### Discontinuing or modifying interventions {11b}

Arms will be discontinued (or modified to remove the risk) if there is evidence that they are harming exposed individuals.

### Adherence {11c}

There are no *a priori* strategies to improve adherence.

### Concomitant interventions {11d}

As our trials will be embedded within routine service delivery, we cannot exclude the possibility that other initiatives will be introduced by local teams before, during, or after individual trials. We will report all programmatic changes that take place during individual trials that could bias our findings.

### Provisions for post-trial care {30}

As this is a negligible risk trial, no provisions will be made for post-trial care.

### Outcomes {12}

#### Primary outcome

The proportion of people attending triage clinic on their appointed date from the left-behind group (adults aged 18-44 years old), measured using attendance data collected by staff when people check-in.

Our left-behind group comprises of younger adults (aged 18-44 years) as this group was found to be the least likely to receive care in a previous study in Meru’s VIP programme.[ref] A focus on left-behind groups is important to programme managers who are trying to close gaps, extend health service coverage, and ensure that their services do not exacerbate existing inequalities.

When referred participants check-in at treatment outreach clinics, attendance status is recorded by administrative staff using the Peek app, which automatically updates a central database that holds records of each participant’s eye care need, sociodemographic characteristics, arm allocation, and attendance status at the ophthalmic clinic on the appointed date. Our Bayesian algorithm will review the attendance data for every referred participant every 7 days and calculate the probability of attendance within each arm. In our modelling we have estimated that 300 people will be referred every 7 days. This aligns with what we have observed in the VIP programme so far, where approximately 1,000 people are screened per day, of whom approximately 1/3 are referred.

#### Secondary outcome

The proportion of people attending triage clinic on their appointed date across the entire population, measured using attendance data collected by staff when people check-in.

If an intervention is found to increase attendance among the left-behind group, we also want to check whether there has been an impact on the overall mean attendance rate. This is to hedge against adopting an intervention that improves access for the left-behind group but leads to a large overall fall in attendance across the entire programme. We will use absolute percentage differences in attendance for comparisons between the left-behind and general populations exposed to the intervention.

### Sample size {13, 14}

As we are using stopping rules, will not pre-specify a minimum sample size or estimate effect sizes for the intervention arms. Instead, participants will be continually recruited until sufficient data accrue to trigger one or more of the other stopping rules. Triallists have argued that this approach is more “efficient, informative and ethical” than traditional fixed-design trials as this approach optimises the use of resources and can minimise the number of participants allocated to ineffective or less effective arms.^17^ Every 7 days the algorithm will review the attendance data and calculate the probability of attendance within each arm.

Based on extensive scenario modelling, we have decided to use the following stopping rules for this trial:

1. There is a >95% probability that one arm is best, i.e. the difference between the two arms is >0%.
2. There is a >95% probability that the difference between the best arm and the arms remaining in the trial is <1%.

### Recruitment {15}

As the trial is pragmatic, the responsibility for recruiting screening participants lies exclusively with local programme managers. Programme implementers will enrol participants by seeking consent from all those who require referral for further assessment and care.

### Allocation

#### Sequence generation {16a}

We will use computer-generated random numbers to generate the allocation sequence and assign all consented, referred participants to intervention arms, with equal numbers of participants in each arm. Where appropriate blocking will be used with blocks between 4-12.

#### Allocation concealment mechanism {16b}

For interventions delivered to individuals, the allocation sequence will be generated within the Peek system in real-time, as participants are referred. As human trial managers are not involved in allocation there is no need for concealment.

### Implementation {16c}

When the random allocation algorithm within the Peek app assigns a patient to the intervention arm, the Peek app will display a notice to the screener that reads ‘Please read script A or B in the patient’s preferred language. The screener will then read the corresponding counselling script from a piece of card (A will be the usual care script, B will be the intervention script). The app will also autogenerate and send the enhanced reminder SMS on the same day, the day before the appointment and on the appointment day to all those assigned to the intervention arm. The control arm will receive the usual SMS reminder on the same day and the day before the appointment.

### Masking

#### Who and how {17a}

Once assigned by the algorithm, each participant’s online record will automatically update to display which arm they have been allocated to. Participants will not be masked to assignment. Screeners will see allocation status as they are required to deliver the intervention. Outcome assessment will be performed by those responsible for checking-in participants at triage clinic. No steps will be taken to mask these staff to participant allocation status. Ongoing interim data analysis will be performed by the Bayesian algorithm every 72 hours.

### Unmasking {17b}

Human investigators and programme managers will not be able to access data on allocation of participants to specific arms unless they are involved in delivering an intervention.

### Data Collection

#### Data collection methods {18a}

Attendance at clinic will be recorded when participants check-in at clinic on their appointed date. Each participant’s attendance status will be recorded on their central record.

### Retention {18b}

There are no plans to promote participant retention and complete follow-up.

### Data management {19, 27}

All data entry will be performed by programme staff as part of routine screening and clinical care. See the data management plan for further information about coding, security, and storage (Additional file 1).

### Statistical methods {20a, 21b, 20b}

All analysis will be conducted using R. Baseline characteristics of all participants will be described as mean (SD) or median (IQR) for categorical variables, or as frequencies and proportions for continuous variables.

During this trial, clinic attendance in each arm will be assessed using Bayesian methods. At each interim analysis point, a binomial distribution of outcome will be described for each arm using the total number of participants allocated to the arm and the number that attended at clinic. The binomial distribution will be combined with a prior distribution to update the posterior distribution of each arm. A regularizing prior of beta(100,100) will be applied to reduce overfitting until a reliable amount of data is accrued. A Monte-Carlo simulation will be used to update posterior distributions at each interim analysis point. Posterior probabilities will be calculated and compared to the stopping rules as to whether the trial should continue into the next day or end early. If there is sufficient evidence to meet one of the stopping rules, the trial will terminate and proceed to the final analysis stage.

Upon completion of the trial, a complete case analysis will be performed on all eligible participants in the trial on an intention-to-treat basis. The primary endpoint of the trial is clinic attendance among the left-behind subgroups after randomization. Within a selected subgroup, the primary analysis will use Monte Carlo simulations to estimate the posterior distribution of attendance between arms. Posterior probabilities will be calculated to compare the proportion of attendance between arms and to identify an arm that results in the highest likelihood of attendance. For the secondary endpoint, analysis will be expanded to all participants in the trial. A more detailed description of the statistical methods will be reported as open access as a separate statistical analysis plan.

### Data Monitoring {5d, 21a}

From UK, Dr Luke Allen, Dr David Macleod (data analyst), Min Kim and Dr Nigel Bolster (PEEK engineer) will have access to all data. In Kenya, Sarah Karanja and Dr Michael Gichangi (Co-Principal investigators) will also have access to these data. Data analysis will be conducted by David and Min Kim and shared with all investigators.

An independent Data and Safety Monitoring Board (DSMB) has been appointed with the primary aim of assuring safety of participants in the trial. The DSMBs will advise the steering committee and sponsor on continuation or stopping of the trial based on safety and efficacy considerations. The DSMB has three members, all independent of the running of the trial, and all with relevant clinical and epidemiological experience. The DSMB will operate independently of the study sponsor and the steering committee. The DSMB will confirm its own specific meeting arrangements and draw up their own charter, working from the template produced by the Damocles Study Group.^18^ It is proposed that the DSMB will meet prior to the beginning of the trial, one third of the way through, and at the end of the trial, to assess the safety of the trial procedures. The DSMB will agree the way it will monitor the data, what it requires from the investigators in this respect and will communicate this to the PIs. All data can be interrogated remotely in real-time. The DSMB may visit the study coordination centre to assess data management, record keeping and other important activities. The DSMB will determine the manner in which it will monitor the data, what it requires from the investigators in this respect and will communicate this to the PIs.

The board comprises a clinical trial specialist who does research in Diabetic Retinopathy, Ophthalmology, Public Health and Health Systems (Dr. Nyawira Mwangi), an ophthalmologist (Dr. Stephen Gichuchi), and a biostatistician (Mr. Moses Mwangi). DSMB will periodically review safety and efficacy data.

### Patient and public involvement

Lay people and a community advisory board has reviewed and contributed to the development of this protocol and all preceding work around identifying the left-behind group and identifying potential service improvements. Lay representatives will assist with interpretation and dissemination of the trial findings.

### Adverse event reporting and harms {22}

An adverse event (AE) is defined as any untoward medical occurrence in a patient or study participant. A serious adverse event (SAE) is defined as any untoward medical occurrence that:

- Results in death
- Is life-threatening
- Requires inpatient hospitalisation or prolongation of existing hospitalisation
- Results in persistent or significant disability/incapacity
- Consists of a congenital anomaly or birth defect

Other ‘important medical events’ may also be considered serious if they jeopardise the participant or require an intervention to prevent one of the above consequences.

All adverse events will be reported. Depending on the nature of the event the reporting procedures below will be followed. Any questions concerning adverse event reporting will be directed to the study coordination centre in the first instance. The flow chart below has been provided to aid the reporting of adverse events.

#### Non-serious AEs

All non-serious AEs will be reported to the study coordination centre and recorded in a dedicated AE log within 72 hours. The entry must state the patient ID, date and time of AE, nature, and relation to the intervention, if any. The AE should also be reported to the data and safety monitoring committee within 72 hours. AE logs will be stored on a secure, password-protected file on a LSHTM computer.

#### Serious AEs

Serious Adverse Events (SAEs) will be reported to the PI and study coordination centre within 24 hours of the local site being made aware of the event (Figure 5). The PI will report the event to the data safety monitoring committee within 48 hours and include it in the study safety report.

**Figure 5:**
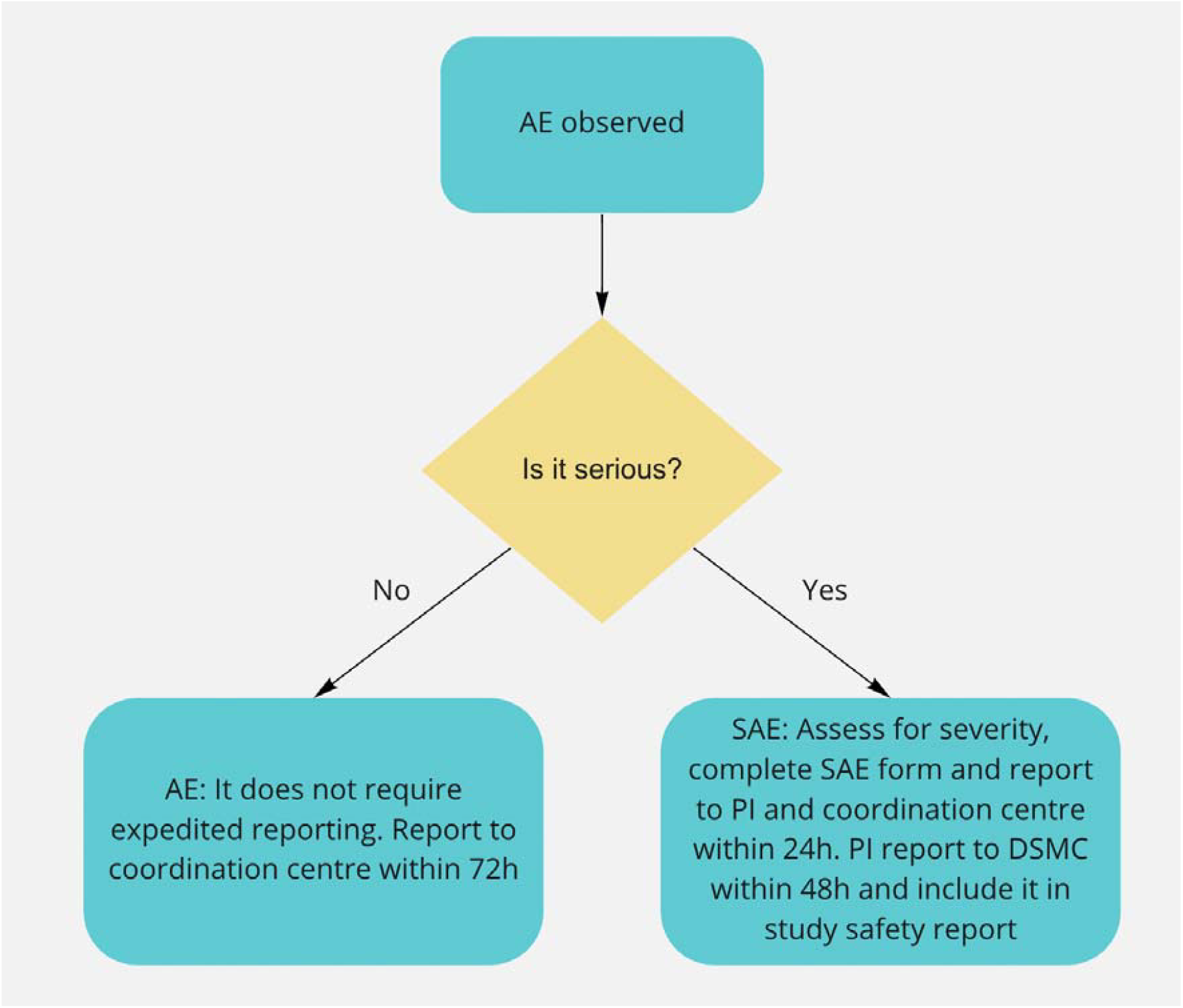
Approach for managing adverse events.

An SAE form will be completed and submitted to the PA and study coordination centre with details of the nature of event, date of onset, severity, corrective therapies given, outcome and causality. All SAEs whether expected, suspected or unexpected will be reported to regulatory bodies and the trial DSMB within 48 hours of occurrence. The responsible investigator will assign the causality of the event. All investigators will be informed of all SAEs occurring throughout the study. If awaiting further details, a follow up SAE report should be submitted promptly upon receipt of any outstanding information.

Any events relating to a pre-existing condition or any planned hospitalisations for elective treatment of a pre-existing condition will not need to be reported as SAEs.

### Contact details for reporting SAEs

SAE forms will be sent to: and the relevant in-country co-PI using the title ‘Urgent - SAE’

### Responsible Personnel

#### Chief Investigator (CI)

- The CI has overall responsibility for the conduct of the study and the ongoing safety and evaluation of any IMPs being used in the trial.
- Promptly notifying all investigators, Institutional Review Board (IRB) or Independent Ethics Committee (IEC) and Competent Authorities (CAs) of each concerned member state of any findings that may affect the health of the trial participants.
- Keeping detailed written reports of all AEs/ARs identified in the protocol as critical to the evaluation of safety within the agreed timeframes specified in the protocol.
- Accurate production and submission of the Development Safety Update Reports and progress reports to CAs and IRB/IECs.
- Collate all AR/AEs/SAEs/SARs and report to the Sponsor annually.
- Ensure that the PIs report all SAEs/SUSARs immediately to the Sponsor and to the CAs, IRB/IECs and any other relevant parties within agreed timelines (
- Supplying the Sponsor and IRB/IEC with any supplementary information they request.

#### Principal Investigators (PI)

- The PIs have responsibility for the research performed at the local site, handling and management of investigational medical products, and informing the CI, Sponsor, Ethics, regulatory bodies and the trial coordinating team, of all adverse events that occur at their site
- Safety responsibilities:
- Ensure trial participant safety and the swift and adequate management of trial participants with any type of AE/AR as per the management protocol described below.
- Reporting all SAEs/SUSARs immediately to the Sponsor and to the CAs, IRB/IECs and any other relevant parties within agreed timelines (i.e. LSHTM, EFMHACA, ORHB, FMOST).
- Assessing each event for causality, severity and expectedness. (Note: a medical decision which must be made by the investigator directly involved with the care of the patient/participant experiencing the AE)
- Ensure adequate archiving of AE records and reports in the local trial office along with the trial master files.
- Collate all AR/AEs/SAEs/SARs biannually and present to the CI.
- Guide and supervise the field research team on accurate recording, reporting of all adverse events.

#### Field Research Team Members (Coordinators, Nurses, Examiners, Recorders)

- All field research team members are responsible for identifying, recording, and reporting any AE or AR to the PIs regardless of severity or causality.
- Assessing each event for causality, severity and expectedness. (Note: a medical decision which must be made by the investigator directly involved with the care of the patient/participant experiencing the AE).
- Ensure that the participant has received the necessary management. This includes advice/reassuring, referral, offering transport, paying for management, making follow-up visits
- Report to the PIs/Project manager AEs/ARs based on the specified timeline and file all AE/AR recorded forms in the trial master file.

### Frequency and plans for auditing trial conduct

The study may be subject audit by the London School of Hygiene & Tropical Medicine under their remit as sponsor, the Study Coordination Centre and other regulatory bodies to ensure adherence to Good Clinical Practice.

## Discussion

### Limitations

It is unlikely that the addition of four items of information will have a large effect size. Nevertheless, the provision of this information was rated as ‘highly likely’ to improve access to clinics by a large majority of those who were surveyed in Meru. This particular intervention is one of many that will be tested in separate trials under the overarching platform trial. Text message reminders have obvious limitations in the context of services for those with poor vision, and many people in Meru do not have their own phone. Every screening participant provides a contact number, and it may be that they can have the message read out to them. Inability to receive or read an SMS message will affect those in the intervention and control arms equally, so this should not introduce bias. With the in-person counselling there is a risk of contamination if screeners end up providing the enhanced counselling information to all participants, irrespective of their allocation. The local trial management team will conduct observations to get a sense of whether this is happening. Contamination would lead to an underestimate of the intervention effect size.

We have chosen to use a prioritarian approach that focuses on left-behind population groups. This prevents a situation where we accept an intervention that improves the overall mean but is associated with a decline among left-behind groups. This approach does not hedge against the slope of inequality worsening. Unfortunately, using a proportionate approach where we assess whether gains in each group are proportionate to their initial need would risk attributing success to our intervention rather than the more likely detection of regression toward the mean.

Our estimate of the probability/proportion will be biased. Because we choose to stop on average at a “local peak”. So for example we’re confident A is better than B, but the estimate of the attendance rate in A will be on average an overestimate.

We use attendance as a proxy for access. Whilst this is the closest hard indicator available, the semantic implication of the term places responsibility on people rather than clinical systems or societal structures. We will counterbalance this in the language that we use to talk about barriers and in the framing of interventions. We also note that we focus on a proximal indicator that does not always correlate well with receipt of high-quality care, or good clinical outcomes. We decided to focus on access for three main reasons; first it aligns with the conceptual narrative of Universal Health Coverage and ‘leaving no one behind’, second attendance data are already routinely collected and available for every single person who is referred, and third, internal Peek data suggests that the ‘fall off’ gap between those who are referred but do not attend is much larger than other gaps e.g. the proportion of those who attend but do not receive appropriate care, or the proportion of those who receive appropriate care but do not experience improved health outcomes.

### Dissemination

The findings will be shared with the programme managers and written up for peer-reviewed publication. No participant names or identifiable information will be used in any of the write-ups. The study findings will be disseminated during quarterly review meetings with implementing partners, community health extension workers and representatives from the county health management committee, and bi-annual partner meetings. We will also publish our findings in peer-reviewed journals, and present abstracts at national, regional and/or international conferences.

## Trial Status

Recruitment has not yet commenced but is planned for March 2024.

## Data Availability

Patient-level data will be pseudo-anonymised removing names and any other key identifiers before it is shared. Only the least amount of data will be shared, and where possible it will be fully anonymised and aggregated. All published findings will be at anonymous aggregate subpopulation level. In line with the UK concordat on open research data (2016), anonymised data from this trial will be made available to bona fide research groups (evidenced via CVs and the involvement of a qualified statistician), and in line with the trials publicly available Data Management Plan (Additional file 1: Appendix 1), following review and approval from the trials data monitoring committee.

## List of Abbreviations

AE: Adverse Events
CI: Chief Investigator
DSMB: Data Safety Monitoring Board
LSHTM: London School of Hygiene and Tropical Medicine
PI: Principal Investigator
RGIO: Research Governance and Integrity Office
SAE: Serious Adverse Event
SMS: Short Message Service
VIP: Vision Impact Project

## Declarations

### Ethics approval and consent to participate {24}

The study was granted ethical approval by the Kenya Medical Research Institute (KEMRI) scientific and ethics review unit, and the London School of Hygiene & Tropical Medicine research ethics committee.

### Consent for publication {32}

Not applicable

### Availability of data and materials {29}

Patient-level data will be pseudo-anonymised removing names and any other key identifiers before it is shared. Only the least amount of data will be shared, and where possible it will be fully anonymised and aggregated. All published findings will be at anonymous aggregate subpopulation level. In line with the UK concordat on open research data (2016), anonymised data from this trial will be made available to bona fide research groups (evidenced via CVs and the involvement of a qualified statistician), and in line with the trial’s publicly available Data Management Plan (Additional file 1: Appendix 1), following review and approval from the trial’s data monitoring committee.

### Trial Sponsor and Contact Information {5b}

London School of Hygiene & Tropical Medicine For further information regarding the sponsorship conditions, please contact the Research Governance and Integrity Office: London School of Hygiene & Tropical Medicine Keppel Street London WC1E 7HT Tel: +44 207 927 2626

### Role of Sponsor and Funders {5c}

The study sponsor and funders will not have any role in- or ultimate authority over the study design; collection, management, analysis, and interpretation of data; writing of the report; or the decision to submit the report for publication.

### Authors contributions {31b}

LA is co-PI. He led the protocol, co-designed the study, and drafted the manuscript. MG is co-PI. He conceived and co-designed the study and revised and approved the manuscript. AB is the chief investigator; he conceived the study, led the funding proposal, and revised and approved the manuscript. DM is the lead statistician. He conceived and co-designed the study, revised and approved the manuscript. NB conceived and co-designed the study, revised and approved the manuscript, and integrated the code into the Peek software. MK conceived and co-designed the study, revised and approved the manuscript, and developed the code. MB conceived and co-designed the study, revised and approved the manuscript. SK developed the intervention and revised and approved the manuscript. MT co-designed the study and read and approved the final manuscript. JC co-designed the study and provided statistical advise on the trial methods. He revised and approved the final manuscript. All author(s) read and approved the final manuscript.

## Acknowledgements

Not applicable

