## Appendix 1 for "Protocol for an individual-level, two arm, superiority RCT within an adaptive platform trial: Enhanced patient counselling and SMS reminder messages to improve access to community-based eye care services in Meru, Kenya"

Appendix 1: Data Management Plan

**1. DATA SOURCES AND DATA COLLECTION PROCESSES**

The research objectives require the collection of quantitative survey data, as well as qualitative data in the form of audio recordings and quotes from study participants. Table 1 below outlines the data fields to be collected throughout the various stages of the data collection process. All data will be treated as personal data for the purpose of data capturing and processing, as collectively, it can be combined in a way that could make it identifiable.

Data from the initial screening process will be collected in Peek powered Eye Health School and Community Programmes using Peek’s Capture application. During the initial screening process only basic and non-personal identifying data is collected, with the exception of telephone number. Following initial screening, all those identified as requiring referral will be asked to provide sociodemographic data to enable us to monitor the equity performance of our programmes e.g. are certain ethnic groups more likely to be screened? The additional sociodemographic indicators are outlined in table 1 below. Based on the visual acuity threshold set prior to screening, the Peek Capture automatically informs the data collector whether the attendee may potentially need onward treatment. For those screened negative no further data is collected. Only for those screened positive is further information collected. This ensures data collection is kept to an absolute minimum maintaining privacy and ensuring compliance with data protection regulations.  For those screened positive, additional information is collected, but the data is always minimised to ensure only the required data is collected at each stage of the service.

Following triage of individuals who had screened positive, a four-stage rapid exploratory sequential mixed-methods study design will be used to evaluate barriers to health access among non-attenders who had been flagged for onward treatment. Telephone interviews will be conducted among 60 non-attenders, purposively selected from socio-demographic groups with the lowest overall attendance rates. The aim of the telephone interviews is to explore and evaluate their perceived barriers to clinic attendance, and develop a list of potential solutions.

Once interventions and service modifications have been identified, these will be tested through a series of pragmatic, embedded, adaptive parallel, multi-arm randomized control trials (APT). The intention of the APT is to continuously improve attendance rates, particularly amongst those groups with the lowest engagement rates overall. Table 1 outlines each of the data collection phases, the data fields to be collected, and the study populations of each of the stages discussed.

Table 1: Data collection phases, data fields and study populations for broader I’M SEEN project

|  | **Phase** | **Data Fields Collected** | **Eligible Population** |
| --- | --- | --- | --- |

| **1.** | **Initial Screening Process** | - Age | - Spectacle status | All included in PEEK screening programme |
| --- | --- | --- | --- | --- |
|  |  | - Gender | - Visual Acuity |  |
|  |  | - Language - Awareness (optional) | - Eye Condition - Telephone Number |  |
|  |  | - Diabetes status (optional) |  |  |
| **2.** | **Collection of sociodemographic data** | - Health insurance status | - Ethnicity | All those identified as requiring referral |
|  |  | - Language | - Disability |  |
|  |  | - Marital Status | - Occupation |  |
|  |  | - Religion | - Education |  |
|  |  | - Migrant/refugee status | - Food adequacy |  |
|  |  | - Housing | - Asset ownership - Family members |  |
| **3.** | **Elicitation questions (via telephone interview)** | *Barrier elicitation questions:*   - In your own words, can you talk me through why we didn’t see you/your child at that clinic?   *Probing questions:*   - Are there any other factors that prevented you/him/her from attending? - Is there anything else you’d like to share? - Of the issues you mentioned, which is the most important? | | Non-attenders of onward treatment appointments purposively selected by sociodemographic group. |
|  |  | *Solution elicitation questions:*  The last part of the interview is exploring whether there is anything we could do to address these barriers and make it more likely that other people like you/children like [child’s name] will attend in the future.   - So, to start, what would make the biggest difference?   *Probing questions:*   - What else would help? - What other changes could we make to the programme that would make it easier for children like [child’s name] to attend? - Are there any other specific changes that we could make to the way that the programme or eye clinics run? - Who do you feel should implement this/these changes?" - You mentioned [list their proposed solutions]. Some of these may be beyond our control, but if we managed to [list their proposed programme-related changes], do you think that would be enough to allow children like your son/daughter to attend?" | |  |

| **4.** | **Online Survey (hyperlink sent via SMS)** | Ranking of proposed service modifications proposed during telephone interview using mobile phone numbers gathered during initial screening process. | Representative sample of non-attenders |
| --- | --- | --- | --- |
| **5.** | **Programme Leader/Stakeholder**  **Workshop** | Audio recording of workshop conversation during which the list of prioritised service modifications derived from the online survey will be discussed and evaluated for testing | Service managers, programme implementers, national and regional eye care policymakers, as well as any other relevant stakeholders. |

| **6.** | **Adaptive Platform Trial** | Examples of possible interventions delivered at the individual and cluster levels include: | | Children over 5 years, and adults who participate in PEEK-powered eye screening programmes. Those who do not meet local clinical service eligibility criteria will be excluded. |
| --- | --- | --- | --- | --- |
|  |  | *Individual* | *Population (cluster)* |  |
|  |  | - SMS messages | - Change to language of messages sent to participants |  |
|  |  | - Voice messages |  |  |
|  |  | - Visual acuity thresholds |  |  |
|  |  | - eVouchers | - Radio broadcasts |  |
|  |  | - Physical vouchers | - Training for implementers |  |
|  |  | - Chaperones | - New clinic times or locations |  |
|  |  | - Individualised transport assistance | - New bus services |  |

**2. DATA COLLECTION TOOLS**

Various data collection tools will be used to populate the data fields outlined in table 1.

*Quantitative Data:*

- Android Mobile Devices – Survey data, and data derived from the APT (phases 1,2, 4 and 6) will be collected by Peek’s implementing partners using Android devices through the Peek Capture application. Peek Capture enforces security controls that include strong device passcodes and native Android encryption. Data stored is time limited, the device syncs via an encrypted connection with a Peek managed server, the data is then deleted to minimise the risk of data stored on the device. The APT will be embedded within Peek software used in parallel with a Bayesian algorithm that will be used to autonomously run response adaptive trials.

*Qualitative Data:*

- Play Verto – The online survey will be administered through Play Verto, a play-based online survey group who have worked with the United Nations and others to develop engaging short surveys that have impressively high response rates in low- and middle-income countries. The survey will be sent as a hyperlink in an SMS. PlayVerto will gather, store and process. After, they will transfer (anonymised data) it to LSHTM who will perform further processing and storage. LSHTM will share aggregate anonymised findings with partners and in public domain.
- Data Abstraction Matrix: During the telephone interviews, data collectors will directly enter notes, quotes, open codes, and abstractions into a matrix. Data gathered, processed and stored by local partner organization. Then shared with LSHTM (fully-anonymised responses to be shared).
- Audio Recordings – Telephone interviews will involve verbal communication and discussion, and thus will be collected and stored using digital audio-recording methods.

**Software:**

- Peek Capture - is an application that runs on Android devices that supports eye health screening and referral pathways to treatment
- Peek Admin - is a web based data platform application that is used to view the data collected by Peek Capture, it tracks the Programme progress, provides insights and helps ensure no one is left behind.
- Play Verto – is a play-based online survey group who have worked with the United Nations and others to develop engaging short surveys that have impressively high response rates in low- and middle-income countries.
- STATA and R, and Excel will be used to analyse the data exported from Peek Admin

**Hardware:**

- Peek servers are hosted on Amazon Elastic Compute cloud-based virtual machines running Amazon Linux.
- Android devices,  locally managed by Peek’s implementing partners.

**3. DATA-RELATED ACTIVITIES**

| **Task** | **Description** |
| --- | --- |
| Start gathering SES data | In month 1 we will start gathering sociodemographic data from:   - a representative sample of all those presenting to be screened - all those identified with an eye care needs and referred on for treatment   These data will be transferred from Android devices in the field to Peek Admin, hosted on AWS.  Note that Peek programmes run continuously and we intend to gather data from participants in every programme so that we can promote equitable service delivery. |
| Clean SES data | Routine manual data cleaning will be conducted periodically by Peek administrators. Internal software guardrails will  pick up simple errors |
| Analyse SES data | Every month we will perform simple descriptive statistical analysis of presentation rates and treatment attendance rates by SES category.  The output of this analysis will be anonymised and presented as mean attendance rates for each SES subgroup e.g. males x%, females z%. |
| Conduct telephone interviews, online surveys and stakeholder workshop | In order to better understand barriers to accessing eye services a series of activities will be conducted through a four-stage sequential mixed-methods approach. These include:  1. Telephone Interviews – Telephone interviews will be conducted with non-attenders, purposively selected from subgroups with the lowest attendance rates.  2. Following telephone interviews, a single list of suggested solutions will be compiled  3. Online survey – An online survey will be conducted among a representative sample of non-attenders to rank mooted interventions/service modifications.  4. Stakeholder workshop – Programme leaders and key stakeholders will then select one or more of the highest ranked interventions to test, based on impact, feasibility, risk and cost.  Following completion of this process, data will be analysed to elicit barriers to care and recommended interventions/service modifications to improve attendance rates. |
| Testing of service modifications through APT | An automated adaptive platform trial (APT) will iteratively test a series of interventions selected with intended service beneficiaries to increases attendance rates among marginalised groups. This will be done through a Bayesian, embedded, pragmatic, superiority, adaptive platform trial platform that will use response adaptive randomisation. |

**Quality checks**

- Errors are flagged at the point of data entry by software that only accepts pre-specified responses e.g. phone numbers must be comprised of a set string length of digits.
- The software has built-in logic steps
- We will institute training and supervision for all data collectors
- Application logging, audit trails and alerting direct administrators to given issues post-collection e.g. when SMS messages fail to be delivered
- Post-collection human data checking using the Peek Admin programme e.g. for ID disambiguation

5. How will you address ethical & legal issues within your research?

- What permissions are needed? E.g. to collect data in country, analyse data for specific purpose, share data
- From whom must approval be obtained? E.g. study participant, ethics committees, data provider
- How will permissions be provided? E.g. ask participants to sign a consent form, sign a Data Transfer Agreement

**4. PERMISSIONS**

Local permissions for Peek powered eye health programmes are already in place. This is in the form of data processing agreements with Peek and the local MoH and/or local implementing partner. This provides a legal agreement between the parties that the data can be collected and processed. The proposed research will be authorised by the same parties to ensure full transparency and the data collection and processing will be managed under the same data processing agreement.

We will obtain written informed consent to collect, analyse, and publish anonymised aggregate participant data in peer-reviewed journals and online open-access data repositories. Individuals will not be identifiable.

In line with UK guidance on risk-adapted approaches to obtaining informed consent, participants will provide consent by ticking a box underneath the following statement:

*“I understand that my anonymous data may be shared with other researchers or online, and that I will not be identifiable from this information. I understand that my decision will not affect the care that I receive, and I am free to change my mind anytime I like.”*

Consent will be obtained when participants initially present for screening.

For screening programmes that include children (<18 years), we will seek consent from their parents/legal guardians using the following statement, sent home on a paper form along with the generic participant information leaflets before screeners visit the school:

*“I understand that my child’s anonymous data may be shared with other researchers. I understand that my child will not be identifiable from this information. I understand that my decision will not affect the care that my child receives, and I am free to change my mind anytime I like.”*

Approval will be sought from research ethics committees at LSHTM and each of the countries where screening takes place.

**5. DOCUMENTATION**

Standard operating procedures and an overall study protocol will be developed in line with LSHTM research guidance to cover all aspects of the research project.

Standardised online training modules have been delivered for programme implementing partners tasked with data collection in the field.

Training will be delivered to all project staff to ensure that they understand the requirements and are able to follow the SOPs.

We have a data compendium which describes the custom sociodemographic variables that we will collect in each country,

**6. DATA STORAGE AND SECURITY**

Data collection, management and storage for this study will be managed by seven entities described below:

A. Peek Vision Capture Application
B. Play Verto
C. The London School of Hygiene and Tropical Medicine
D. Botswana: The University of Botswana
E. India: Dr Shroff Charity Eye Hospital
F. Kenya: Kenya Medical Research Institute?
G. Nepal: Nepal Netra Jyoti Sangh

**Peek Capture Application**

**Pre research data collection and storage in Peek powered eye health programmes**

The data will be collected in Peek powered Eye Health School and Community Programmes using Peek’s Capture application.  Data will be collected by Peek’s implementing partners using Android devices through the Peek Capture application. Peek Capture enforces security controls that include strong device passcodes and native Android encryption. Data stored is time limited, the device syncs via an encrypted connection with a Peek managed server, the data is then deleted to minimise the risk of data stored on the device.  h

The data is stored on a Peek managed server hosted in a Virtual Private Cloud (VPC) utilising the Amazon Web Services (AWS) Cloud. Each Peek powered programme is hosted on it’s own dedicated server and a VPC that will reside in the UK/EU ensuring all of the data privacy safeguards as governed under the GDPR. All data collected is securely stored in AWS data centers which are state of the art, utilising innovative architectural and engineering approaches.  More information, including a virtual tour, can be found by visiting the link [here](https://aws.amazon.com/compliance/data-center/).

Throughout the eye health programme life cycle only approved implementation partners and Peek team members have access to programme data. Access is strictly controlled through the Peek Admin web based data platform application. This is used to view the data collected by Peek Capture, it tracks the Programme progress, provides insights and helps ensure no one is left behind.

**Peek Capture security:**

- Peek Capture is installed on implementing partners managed Android devices
- Peek Capture enforces security controls that include strong device passcodes and native Android encryption.
- Data stored is time limited, the device syncs via an encrypted connection with a Peek managed server, the data is then deleted to minimise the risk of data stored on the device.

**Peek Admin security:**

- Strong passwords, minimum of 12 characters, password strength meter where only ‘strong’ is accepted, blacklist passwords are enforced to ensure easily guessed and passwords found in data breaches cannot be used.
- 2-Factor Authentication to protect user account security.
- User access permissions are controlled through account privileges, this controls scope of programme so access is restricted and limited to only what a user requires for their work, admin privileges are restricted to only those that require the access, account management and patient level reporting.
- Accounts disable automatically after 60 days of inactivity.
- User access reviews available for implementing partners to ensure leavers and inactive accounts are removed.

**Peek Platform Data Security Assurance:**

Peek is an International Standardisation Organisation (ISO) 27001 certified organisation. ISO 27001 certification requires an annual audit by an accredited external auditing body who verify compliance with the industry best practice information security controls.

Peek servers hosted in a Virtual Private Cloud (VPC) utilising the Amazon Web Services (AWS) Cloud. Each Peek powered programme is hosted on it’s own dedicated server and a VPC that will reside in the UK/EU ensuring all of the data privacy safeguards as governed under the GDPR. All data collected is securely stored in AWS data centers which are state of the art, utilising innovative architectural and engineering approaches.

More information, including a virtual tour, can be found by visiting the link below:

<https://aws.amazon.com/compliance/data-center/>.

Annual penetration tests conducted by a 3rd party specialist security testing company. The purpose of the test is to verify whether robust security mechanisms are in place to prevent unauthorised users from accessing data and infrastructure. This penetration test includes:

- Identification of potential vulnerabilities occurring in the application and defining possible attack scenarios conducted with techniques typical for attacks on web applications;
- Simulated attacks from the perspective of an anonymous and standard user;
- Testing API endpoints from the perspective of an anonymous and standard user, including mechanisms such as user authentication, access control, and data validation;
- Security assessment of our infrastructure against the latest industry standard AWS CIS Foundations Benchmark.

The AWS Compliance Program provides further assurance and understanding of the robust controls in place to maintain security and compliance in the cloud. AWS regularly achieves third-party validation for thousands of global compliance requirements that are continuously monitored to meet security and compliance standards for the most sensitive data and privacy requirements. AWS supports more security standards and compliance certifications than any other offering, including PCI-DSS, HIPAA/HITECH, FedRAMP, GDPR, FIPS 140-2, and NIST 800-171, helping satisfy compliance requirements for virtually every regulatory agency around the globe. More information can be found by visiting <https://aws.amazon.com/compliance/programs/>.

**Peek Platform Data Security Controls:**

*Peek Servers:*

Peek servers hosted in a Virtual Private Cloud (VPC) utilising the Amazon Web Services (AWS) Cloud. Each Peek powered programme is hosted on it’s own dedicated server and a VPC that will reside in the UK/EU ensuring all of the data privacy safeguards as governed under the GDPR.

Server OS is Amazon Linux ustlising AWS AMIS to provide base images for our system drives and enhances security by focusing on two main security goals, limiting access and reducing software vulnerabilities. Security updates are applied automatically to test once a week and then rolled out a week later automatically to other environments

*Docker:*

Peek server software runs in Docker containers. Docker shields application software from variations in platform and co-hosted software. It ensures that development, test and production environments run the same context as one another to ensure consistent, predictable behaviour. Peek servers also use docker swarm mode to achieve failsafe reliability and replication of Mongo databases.

*Databases:*

Server data is stored in Mongo databases, a fast, scalable, json document database. Peek infrastructure uses a Mongo replica set across two hosts. There are two replicas each holding a full copy of the data and one arbiter. The arbiter is only used for the election of a new master if one of the nodes was to become unavailable. The Mongo database and journal are held on AWS Secure EBS volumes. This provides 256-bit AES encrypted using a key managed under the Amazon Key Management Service.

Amazon Key Management Service, allows us to create and manage cryptographic keys and securely control their use across a wide range of AWS services and within our applications. AWS KMS is a secure and resilient service that uses hardware security modules that have been validated under FIPS 140-2 to protect the encryption keys. AWS KMS also integrates with AWS CloudTrail providing us with secure logs of all key usage. Backups on S3 are also encrypted using keys managed by AWS Key Management Service.

*Logging and Monitoring:*

Peek Server and Mongo Server logs and uploaded to AWS Cloudwatch for storage and monitoring. AWS Cloudwatch collects monitoring and operational data in the form of logs, metrics, and events and alerts us immediately of problems in any environment, both application and infrastructure.

*Network Security:*

AWS Security groups are used to provide firewall-like network access control and allow inbound traffic on HTTP and  HTTPS ports. Outbound traffic is permitted on any port. The SSH traffic is restricted to subnets associated with devops engineers and the deployment servers. TLS 1.2 is used to secure traffic between device or browser and server.

Operational access to the AWS console is protected with AWS IAM MFA which uses 2-Factor Authentication and ensures that access to AWS is restricted to users with knowledge of password and possession of a specific approved mobile device. Automated access to the AWS API uses AWS Roles with restricted privileges needed for housekeeping, logging and alarm maintenance. No user use is made of Access Keys to eliminate the vulnerabilities of file-system-based credentials.

*Threat Detection:*

AWS Guard Duty is enabled, this provides  a threat detection service that continuously monitors for malicious activity and unauthorised behaviour to protect access, workloads and data. The service utilises up-to-date threat intelligence feeds from AWS, CrowdStrike, and Proofpoint and continuously evolves through machine learning.

*Backups:*

An Image is maintained of the Server Host using AWS AMI to ensure continuous availability.

A snapshot of the encrypted data volume, containing database and journal, is taken four times daily. Snapshots are retained for two weeks. Access to the snapshots is strictly controlled. Old backups are automatically deleted after 90 days. Backups are stored on AWS S3 storage, also encrypted providing 256-bit AES encryption. The backups are stored across AWS multiple availability zones, this ensures that the data resides in multiple data centres separated geographically and stored in AWS secure data centres.

Additionally, a further backup is made off AWS. Off-AWS backups are replicated to Google Cloud daily via Google Transfer service to identically named buckets and files with a retention policy of 90 days.

*Data Centres:*

All data collected is securely stored in AWS data centers which are state of the art, utilising innovative architectural and engineering approaches.

*Disaster Recovery:*

A full disaster recovery test is performed at least annually to ensure servers, applications and databases can be fully recovered within 24 hours.

**Play Verto**

*Play Verto Data capture tool*

Data collection via our web-based application is all stored on a AWS RDS dedicated server, located in Ireland. This database utilises AWSs own encryption, AES-256 at rest, for maximum security. All data collected is securely stored in AWS data centers which are state of the art, utilising innovative architectural and engineering approaches.  More information, including a virtual tour, can be found by visiting the link [here](https://protect-eu.mimecast.com/s/fPr8C4LO0CEW0E6SO_dRC?domain=aws.amazon.com).

Only approved team members have access to the data. Access is strictly controlled through the Play Verto’s Admin and AWS Admin. Where Password protection is required and the use of 2-factor authentication where applicable.

*Play Verto Capture security:*

- Play Verto is a web-based application therefore can only be accessed via a public URL.
- Play Verto enforces security controls that include strong device passcodes and 2-factor authentication where applicable…
- Data stored is encrypted via AES-256 encryption

*Play Verto  Admin security:*

- We have a strong password policy in place for all our accounts, requiring a minimum length of 8 characters.
- 2-Factor Authentication to protect user account security.
- User access permissions are controlled through account privileges. So access is restricted and limited to only what a user requires for their work.

*Play Verto Platform Data Security Assurance:*

Play Verto  complies with CyberEssentials Certification and IASME Governance Standard. Data collection via our web-based application is all stored on a AWS RDS dedicated server, located in Ireland. This database utilises AWSs own encryption, AES-256 at rest.

Monthly automated penetration tests conducted by [Detectify](https://protect-eu.mimecast.com/s/-jvsC5LPGC3Kj3RHO3UYA?domain=detectify.com" \t "_blank) The purpose of the test is to verify whether robust security mechanisms are in place to prevent unauthorised users from accessing data and infrastructure. We have maintain Threat score of 0 and 10/10, OSWASP SCORE *(The worldwide non-profit organization Open Web Application Security Project (OWASP)’s list of the ten most common vulnerabilities, known as OWASP Top 10, is often used as a security standard. Detectify covers OWASP Top 10 and provides an easy way for you to see which categories you pass or fail.)*

The AWS Compliance Program provides further assurance and understanding of the robust controls in place to maintain security and compliance in the cloud. AWS regularly achieves third-party validation for thousands of global compliance requirements that are continuously monitored to meet security and compliance standards for the most sensitive data and privacy requirements. AWS supports more security standards and compliance certifications than any other offering, including PCI-DSS, HIPAA/HITECH, FedRAMP, GDPR, FIPS 140-2, and NIST 800-171, helping satisfy compliance requirements for virtually every regulatory agency around the globe. More information can be found by visiting [https://aws.amazon.com/compliance/programs/](https://protect-eu.mimecast.com/s/QYbzC69QJS79O7MSmh3Vo?domain=aws.amazon.com).

**Play Verto Platform Data Security Controls:**

*Play Verto  Servers:*

Data collection via our web-based application is all stored on a AWS RDS dedicated server, located in Ireland. This database utilises AWSs own encryption, AES-256 at rest, for maximum security. Ensuring all of the data privacy safeguards as governed under the GDPR.

*Databases:*

Server data is stored in Mongo databases, a fast, scalable, json document database. Play Verto infrastructure uses a Mongo replica set across two hosts. There are two replicas each holding a full copy of the data and one arbiter. The arbiter is only used for the election of a new master if one of the nodes was to become unavailable. The Mongo database and journal are held on AWS Secure EBS volumes. This provides 256-bit AES encrypted using a key managed under the Amazon Key Management Service.

Amazon Key Management Service, allows us to create and manage cryptographic keys and securely control their use across a wide range of AWS services and within our applications. AWS KMS is a secure and resilient service that uses hardware security modules that have been validated under FIPS 140-2 to protect the encryption keys. AWS KMS also integrates with AWS CloudTrail providing us with secure logs of all key usage. Backups on S3 are also encrypted using keys managed by AWS Key Management Service.

*Logging and Monitoring:*

Play Verto Server and Mongo Server logs and uploaded to AWS Cloudwatch for storage and monitoring. AWS Cloudwatch collects monitoring and operational data in the form of logs, metrics, and events and alerts us immediately of problems in any environment, both application and infrastructure.

*Network Security:*

AWS Security groups are used to provide firewall-like network access control and allow inbound traffic on HTTP and  HTTPS ports. Outbound traffic is permitted on any port. The SSH traffic is restricted to subnets associated with devops engineers and the deployment servers. TLS 1.2 is used to secure traffic between device or browser and server.

Operational access to the AWS console is protected with AWS IAM MFA which uses 2-Factor Authentication and ensures that access to AWS is restricted to users with knowledge of password and possession of a specific approved mobile device. Automated access to the AWS API uses AWS Roles with restricted privileges needed for housekeeping, logging and alarm maintenance. No user use is made of Access Keys to eliminate the vulnerabilities of file-system-based credentials.

*Threat Detection:*

AWS Guard Duty is enabled, this provides  a threat detection service that continuously monitors for malicious activity and unauthorised behaviour to protect access, workloads and data. The service utilises up-to-date threat intelligence feeds from AWS, CrowdStrike, and Proofpoint and continuously evolves through machine learning. 

*Backups:*

An Image is maintained of the Server Host using AWS AMI to ensure continuous availability.

A snapshot of the encrypted data volume, containing database and journal, is taken four times daily. Snapshots are retained for two weeks. Access to the snapshots is strictly controlled. Old backups are automatically deleted after 90 days. Backups are stored on AWS S3 storage, also encrypted providing 256-bit AES encryption. The backups are stored across AWS multiple availability zones, this ensures that the data resides in multiple data centres separated geographically and stored in AWS secure data centres.   

Additionally, a further backup is made off AWS. Off-AWS backups are replicated to Google Cloud daily via Google Transfer service to identically named buckets and files with a retention policy of 90 days.

*Data Centres:*

All data collected is securely stored in AWS data centres which are state of the art, utilising innovative architectural and engineering approaches.

*Disaster Recovery:*

A full disaster recovery test is performed at least annually to ensure servers, applications and databases can be fully recovered within 24 hours.

-----------------------------------------

**EXPORT DATA SHARING FOR ANALYSIS** At the analysis stage pseudo-anonymised data will be exported in an encrypted zip file CSV file to LSHTM researchers to perform statistical testing. The zip file will be saved on the protected LSHTM server and only named project staff will be given access. Passwords will be sent separately. We will only ever export the minimum data required for the analyses.

**Labelling conventions**

1. Keep file names short, meaningful and easily understandable to others.

2. Order the elements in a file name in the most appropriate way to retrieve the record.

3. Avoid unnecessary repetition and redundancy in file names and paths

4. Avoid obscure abbreviations and acronyms. Use agreed University abbreviations and codes where relevant.

5. Avoid vague, unhelpful terms such as “miscellaneous” or “general” or “my files”

6. Use capital letters to delimit words, as the preferred option, although underscores (_) or hyphens (-) may add clarity, they make the file name longer.

7. For numbers 0-9, always use a minimum of two digit numbers to ensure correct numerical order (e.g. 01, 02, 03 etc.)

8. Dates should always follow same format: YYYY-MM-DD e.g. 2017-04-25

9. When including a personal name give the family name first followed by initials, with no comma in between e.g. SmithAB

10. Avoid using common words such as ‘draft’ or ‘letter’ at the start of file names unless doing so will make it easier to retrieve the record.

11. Use alphanumeric characters i.e. letters (A-Z) and numbers (0-9). Avoid using invalid characters in file names such as *? \ / : # % ~ { }

12. The file names of records relating to recurring events should include the date and a description of the event, except where the inclusion of these elements would be incompatible with rule 3.

13. The version number of a record should be indicated in its file name by the inclusion of ‘V’ followed by the version number (e.g. V01, V03 etc.). However versioning is enabled automatically in systems such as Office 365 and One Drive for Business, making it unnecessary to duplicate this information in the file name itself.

e.g. 2021-11-19_Topic_Filename-variable01

How will we keep data safe and secure?

- Delete personal & confidential details at the earliest opportunity (specify when)
- Use digital storage that require a username/password or other security feature
- Physical security (such as locked cabinet or room)
- Encrypt storage devices
- Encrypt data during transfer
- Avoid cloud services located outside EU
- Take ‘Information Security Awareness training’
- Ensure backups are also held securely

The aggregated data that is shared among project staff and partners will not contain any names, however the data being shared may still permit the identification of individuals depending on the domains being shared and may therefore constitute pseudo-anonymised data.

We also note that there is not adequate shared secure storage space at LSHTM. We will have to use our personal H drives which is suboptimal for joint working and version control.

**ARCHIVING & SHARING**

All data will be stored for 10 years.

- Files intended for sharing may be hosted in the LSHTM data repository ([http://datacompass.lshtm.ac.uk](http://datacompass.lshtm.ac.uk/)) or a 3rd party repository, such as UK Data Service, ArrayExpress, Zenodo, etc.
- Internal and confidential files can be held on the LSHTM Secure Server
- Internal confidential files will be retained on Peek’s secure servers.
- LSHTM analyses will be saved on encrypted and password-protected files on LSHTM SharePoint, with access restricted to the project team. Once the project is complete these files will be moved to a secure server.
- Data presented in publications (anonymised aggregate mean attendance rates for each SES subgroup) will be published on GitHub.

Resources will be made available at the same time as findings are published in an academic journal. Once available, we will make other researchers aware that the resources exist by:

- Citing resources in future research papers, e.g. in the data access statement or reference list
- Citing resources in project reports
- Adding resources to a list of our academic outputs

The following steps will be taken to ensure that resources are easy to analyse and use in future research:

- Store resources in open file formats such as CSV, Rich Text, etc. See <https://www.ukdataservice.ac.uk/manage-data/format/recommended-formats>
- Designate a corresponding author / data custodian who will handle data-related questions

Conditions on access/use

| Requirement: | To be addressed by: |
| --- | --- |
| In line with the UK concordat on open research data (2016), anonymised data from this trial will be made available to bona fide research groups (evidenced via CVs and the involvement of a qualified statistician), and in line with the trial’s publicly available data sharing policy, following review and approval from the trial’s data monitoring committee. No reasonable request will be turned down, and the appropriate data will be made available within 1-month of receiving the request.    There may be multiple levels of permission required in-country before data can be shared, including national ministry of health approval and local implementation partner approval | The PI will forward requests for data to the in-country leads in order to seek the relevant permissions. We will respond to any boa fide request within 28 days. |

**RESOURCING**

With respect to costs of resources, we have adequate funding within the Wellcome project grant. The data is collected through active live Peek powered programmes where funding and resources is already provided for data collection and data security.
